# Recent measles virus infection increased the severity of infectious disease in WWI with the exception of pandemic influenza

**DOI:** 10.64898/2026.03.16.26348545

**Authors:** Lauren E. Steele, Melanie Wu, Jane E. Sinclair, Kirsten Ignacio, Kyle L. Macauslane, Georgina McCallum, Ellesandra C. Noye, Katina D. Hulme, Nathalie A. J. Verzale, Isabelle Hocking, Megan Airey, Sevim Mese, Michael Waller, Svenn-Erik Mamelund, Carolien E. van de Sandt, Keng Yih Chew, Meagan Carney, Kirsty R. Short

**Author notes:** **Corresponding author:** Kirsty R. Short, Postal address: School of Chemistry and Molecular Biosciences, Chemistry Building 68, Cooper Road, The University of Queensland, St Lucia, 4072, Queensland, Australia. These authors contributed equally to this work (Carolien E. van de Sandt, Keng Yih Chew, Meagan Carney, Kirsty R. Short).

## Abstract

**Background:** In World War 1 (WW1) outbreaks of measles were associated with high case fatality rates amongst soldiers. Recent studies have shown that survivors of acute measles can also develop immune amnesia, increasing their susceptibility to other infections. However, the impact of prior measles infection on infectious diseases during WWI remains unclear.

**Methods:** Here, we create a searchable database documenting the medical history of 1,569 individuals from the Australian, New Zealand, and Canadian forces during WW1.

**Results:** We use this novel database to show that a recent measles hospitalisation was associated with a higher chance of death for infectious diseases (excluding pandemic influenza like illness), consistent with immune amnesia. Surprisingly, a prior measles infection was associated with a significant reduction in hospitalisations duration from pandemic influenza like illness.

**Conclusion:** These findings highlight the unique interaction between measles and pandemic influenza, contrasting with other infectious diseases, and underscore the significant health burden measles placed on young adults during WW1.

## INTRODUCTION

Measles is a highly contagious viral disease that can cause severe complications, including pneumonia, encephalitis and death. For individuals fortunate enough to survive the acute measles infection, there are also long-term immunological consequences. Measles virus induces what is referred to as ‘immune amnesia’ by depleting the pre-existing acquired immunological memory to other pathogens^1^. While leukopenia tends to be transient, individuals show increased susceptibility to secondary infections years after the initial measles virus infection^1,3,4^. Thankfully, since the 1960s an effective measles vaccine has been available, and it is estimated in 2023 that 83% of children globally have received at least one dose of this vaccine. Accordingly, at present only select populations remain unvaccinated, making it difficult to study measles-induced susceptibility to other infectious diseases in real time on a large scale. Whilst some such studies have been performed in unvaccinated, religious communities^5^ a historical analysis of measles cases prior to the widespread availability of the measles vaccine has the potential to afford new insights into measles pathogenesis.

Measles-associated illness was a significant problem for individuals drafted during WW1 (1914-1918). Men recruited from remote and rural areas who may have had limited exposure to many childhood diseases, including measles, were at particular risk of infection^6-8^. Case fatality rates for measles in recruits could be up to 12%, as in certain divisions of the British Army^6^. Indeed, a US army-wide measles outbreak in 1917 resulted in more than 95,000 cases and more than 3,000 deaths^7^. Numerous other infectious diseases were also prevalent during WWI. It is estimated that approximately 10% of deaths in the German army were the result of disease^10^. Common diseases included tuberculosis, malaria, venereal disease and trench fever^10,11^. Of course, WWI also partly coincided with the 1918 influenza pandemic, which killed 40-100 million people worldwide.

Military health records thus represent a unique resource to examine the effects of a prior measles virus on the severity of subsequent infectious diseases in WWI, including 1918 influenza. However, the use of such records is challenging. The detailed medical records of Allied enlisted individuals can be obtained from the National Archives of Australia^16^, the Archives of New Zealand^17^ and The Library and Archives Canada^18^. However, this frequently requires reading multiple pages of scanned, handwritten documentation, the format of which is inconsistent between different individuals. Here, we develop a searchable and validated database combining personnel records from the Australian Imperial Force, the New Zealand Expeditionary Force, and the Canadian Expeditionary Force from the 8^th^ of February 1915 to the 5^th^ of April 1922. We use this combined database to provide the first evidence that a recent measles infection was associated with an increased risk of death with infectious disease in WWI, with the notable exception of 1918 pandemic influenza like illness (ILI). In contrast, a prior measles virus infection was associated with a significant reduction in the severity of pandemic ILI.

## METHODS

### Australian Imperial Force (AIF) and New Zealand Expeditionary Force (NZEF)

The AIF project database^21^ and the NZEF project database^22^ were searched in their entirety to screen for individuals who were continuously enlisted between 1/1/16 to 31/12/18. This was to reflect the fact that measles can induce immunosuppression for up to three years after the acute infection^1,3,4^. The exception to this was if they died of disease during the enlistment period, then they were still included in the database if they were enlisted in the three years prior to death. Of the 257,607 Australian records searched, 1,146 (0.44 %) fit this inclusion criteria and had a direct link to personnel records maintained in the National Archives of Australia^18^ (Supplementary Figure 1A). Of the 3,338 New Zealand records searched, 216 (6.47%) fit this inclusion criteria and had a direct link to personnel records maintained in the Archives New Zealand^19^ (Supplementary Figure 1B). Occupations were assigned as either blue collar or white collar according to a consensus of three independent co-authors (Supplementary Table 1). Rurality was assigned as either rural or urban according to the population of soldier location of birth, as specified by the Australian 1991 Census Edition. Populations were obtained from online published 1911 census. Populations <10,000 were defined as rural. Complexion was categorised as ‘light’ or ‘dark’ by three independent co-authors (Supplementary Table 2). The date of hospitalisation for disease was recorded as the date of admission to hospital or field ambulance. ILI was inclusive of acute bronchitis, acute bronchitis pleurisy, bronchial pneumonia, bronchitis, bronchopneumonia, double bronchopneumonia, grippe, la grippe, double lobar pneumonia, double pneumonia, influenza, influenza pneumonia, lobar pneumonia, pneumonia toxaemia, septic pneumonia, pleurisy with effusion, pleural effusion, pleurisy, pleuro/lobar pneumonia, pneumonia, and septic bronchopneumonia. Influenza was defined as seasonal (ILI before April 1918) or pandemic (ILI from April 1918 to December 1919, inclusive).

### Canadian Expeditionary Force (CEF)

Military hospital admission records from military hospital admission and discharge books from military districts 1-3 in Ontario, Canada^1^ were searched to screen for individuals who were hospitalised with influenza or measles during military service. Of the approximately 54,850 Canadian records searched, 207 (0.38%) fit this inclusion criteria and had a direct link to personnel records from The Library and Archives Canada, First World War Personnel Records^20^ (Supplementary Figure 1C). Individuals were only included in the database if they were continuously enlisted for a period of 3 years between 08/02/1915 and 05/04/1922. The exception to this was if they died of disease during the enlistment period, then they were still included in the database if they were enlisted in the three years prior to death. Military hospital records of these individuals were obtained from The Library and Archives Canada, First World War Personnel Records database using their regimental number and names. Data was then recorded as described from the AIF and NZEF.

### Statistical analysis and data availability

Only individuals with non-missing relevant data were used in the regression model and individuals surviving long enough to be exposed to 1918 pandemic influenza (e.g. individuals surviving to at least 1 July 1918) were included for multiple linear regression modelling.

After exclusion, a total of 1,306 individuals remained to be used in regression analysis, of those, 54 had a history of measles infection. To investigate the effect of explanatory variables on an individual’s severity outcome of pandemic influenza, an outcome variable “days in hospital” was calculated using the difference in days between the available admission and discharge dates. Multiple linear regression was then performed by statistically fitting the intercept *β*_0_ and coefficients *β*_1_, … *β*_*n*_, to best approximate the relationship between the explanatory variables and the outcome variable in the model,

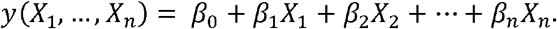

Power analyses were performed post statistical fitting for the coefficient associated to the lowest group size (measles). A required 49 individuals are found to be needed to observe statistical significance at the *α* = 0.05 confidence level with a statistical power of 70%. Not surprisingly, this result indicates that we can expect to observe statistical significance of the coefficient found for the measles grouping.

All statistical analyses were performed in MATLAB (R2022a). The database created herein and code can be found using the DOI: https://doi.org/10.48610/f7e4c44.

## RESULTS

### A validated database of AIF, NZEF and CEF medical records

We identified 1,146 Australian individuals, 216 New Zealand individuals and 207 Canadian individuals who met the predefined study criteria (Supplementary Figure 1). The collection of these data required the interpretation of >300,000 often handwritten, records (Figure 1A). Accordingly, numerous potential sources of error in the construction of the data were identified including errors in original bookkeeping, errors on source website and data translation errors (Supplementary Figure 2). To provide some insight as to the accuracy of the database the distribution of two of the collected, continuous variables (body mass index [BMI] and age) were assessed (Figure 1B & C). As expected in a military population in WWI, the distribution of enlistment age showed that the majority of individuals (99.41%) were below 45 years of age (Figure 1B), consistent with the limit of 45 years for enrolment in the WW1 Imperial and Expeditionary Forces^18-20^. A BMI in the obese range (BMI ≥ 30 kg/m^2^) was uncommon in males in the early 1900s^21^. This fact is reflected in the BMI distribution of the individuals in the collated database where few (4/1528) were recorded as having a BMI ≥ 30 kg/m^2^ (Figure 1C). To ascertain the accuracy of these records, further information was sought about individuals with a BMI ≥ 30 kg/m^2^. An Australian individual with a BMI of 35.8 was a nurse. Upon researching her name, a photograph of her was found, and it was determined that her recorded height and weight were likely to be correct. This is consistent with diversity of body types reported elsewhere amongst Australian nurses in WWI (Figure 1D). Together, these data suggest that the distribution of continuous variables in the database was consistent with what would be expected with this population group.

**Figure 1:**
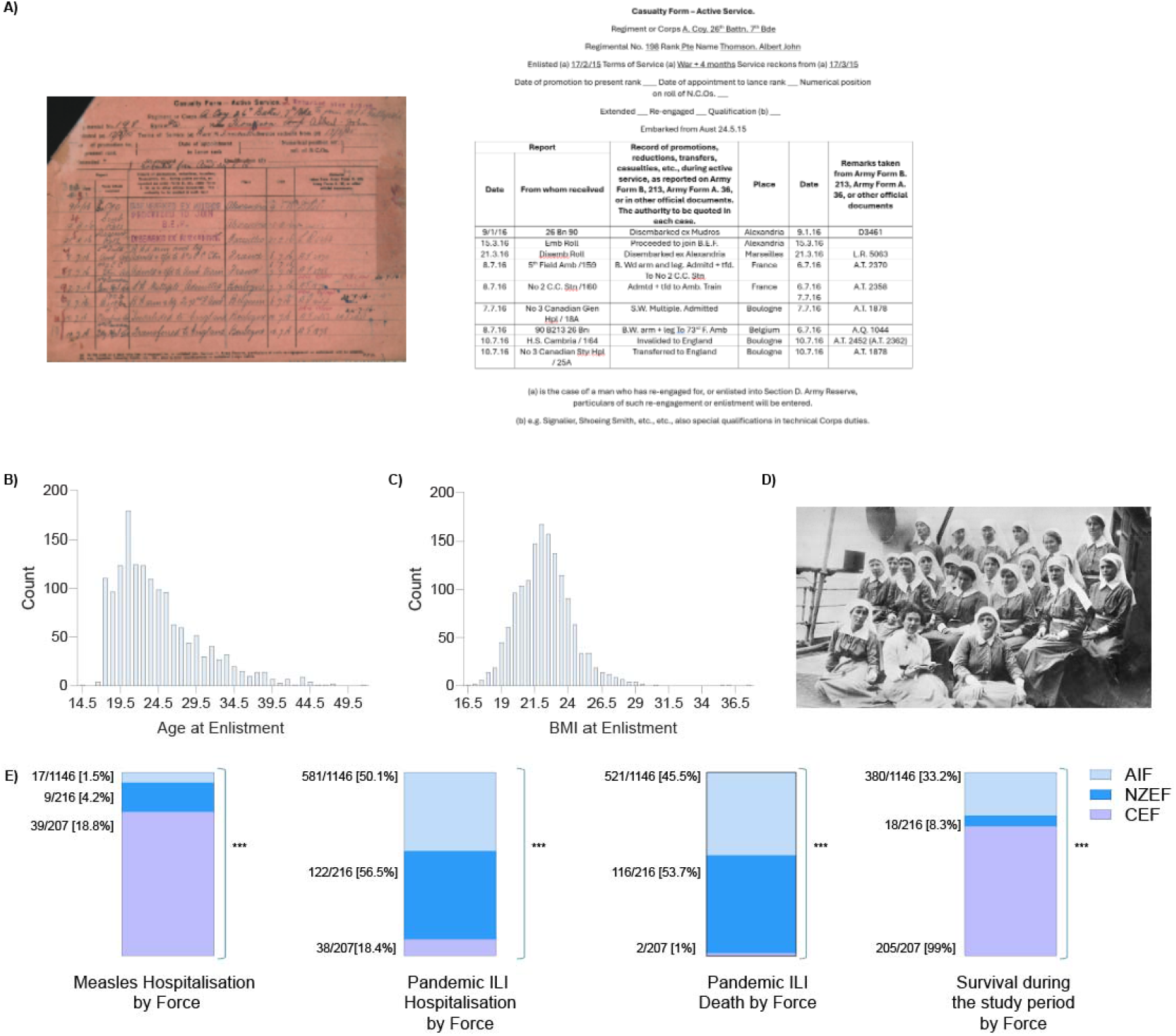
Database characterisation. **A**) Representative image of handwritten casualty form from the AEF and the corresponding digitised information. The distribution of age **(B**) and BMI (**C**) amongst individuals in the database. (**D**) Photo of Australian nurses serving in WWI. Image sourced from https://www.abc.net.au/news/2014-08-04/world-war-i-australian-nurses-missed-out-on-recognition/5642274. (**E)** Force dependence differences in measles incidence, pandemic ILI hospitalisation, pandemic ILI death and survival during the study period. Statistical significance was determined using a Chi-square test. ***p<0.01. (**B, C & E**) n = 1528.

To provide further insight into the strengths and limitations of the collated database the number of missing values were assessed across all forces in the categories of age, sex, complexion, enlistment date, BMI, occupation and rurality. Across all three forces there were no missing values for age, sex or enlistment date. All other missing values constituted <3% of the dataset, except for rurality (7.3%) (Table 1).

**Table 1:**
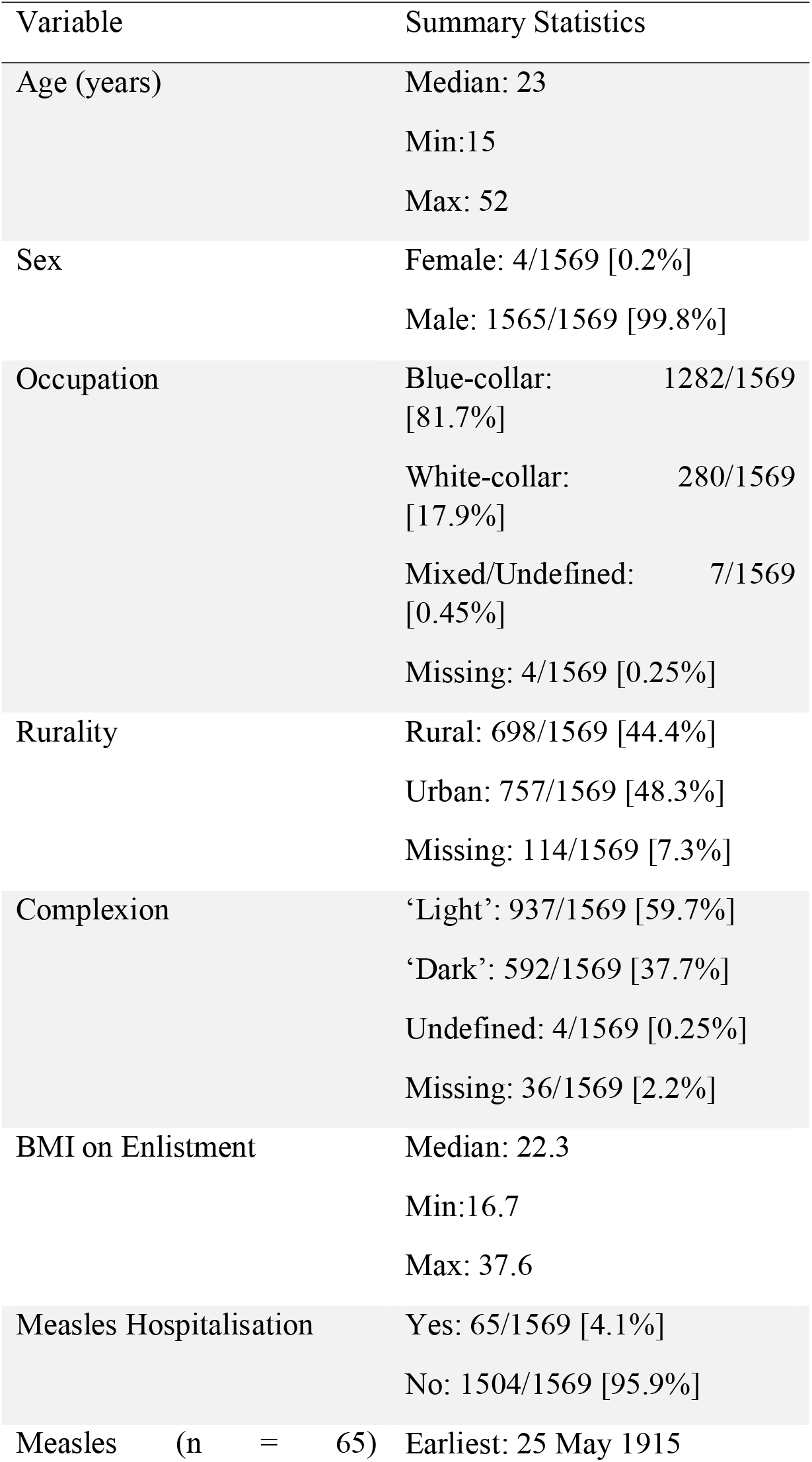
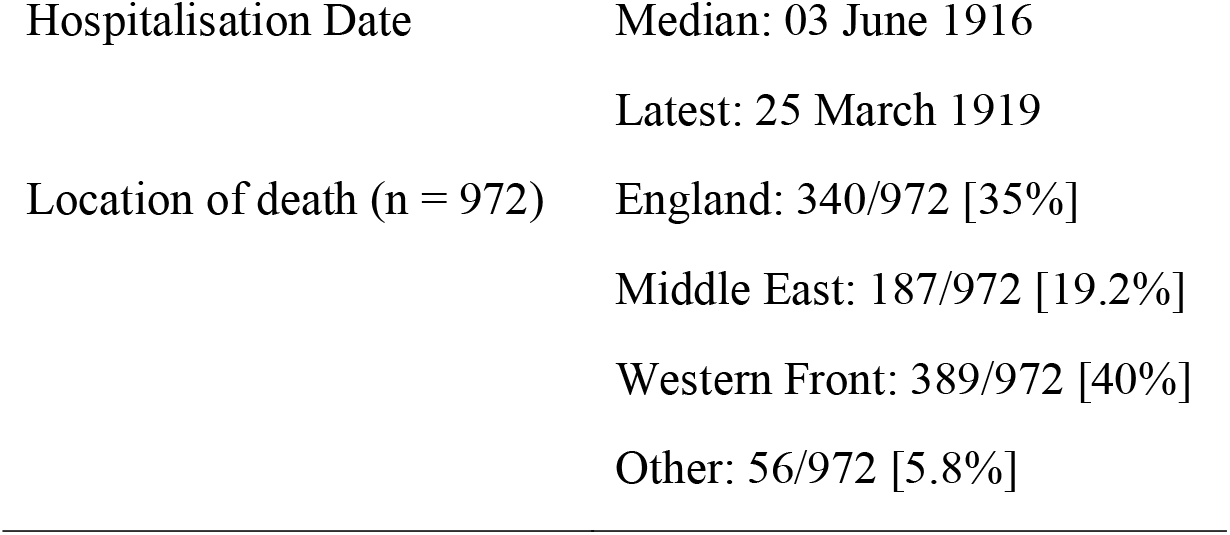
Military personnel characteristics.

| Variable | Summary Statistics |
| --- | --- |
| Age (years) | Median: 23<br>Min: 15<br>Max: 52 |
| Sex | Female: 4/1569 [0.2%]<br>Male: 1565/1569 [99.8%] |
| Occupation | Blue-collar: 1282/1569 [81.7%]<br>White-collar: 280/1569 [17.9%]<br>Mixed/Undefined: 7/1569 [0.45%]<br>Missing: 4/1569 [0.25%] |
| Rurality | Rural: 698/1569 [44.4%]<br>Urban: 757/1569 [48.3%]<br>Missing: 114/1569 [7.3%] |
| Complexion | ‘Light’: 937/1569 [59.7%]<br>‘Dark’: 592/1569 [37.7%]<br>Undefined: 4/1569 [0.25%]<br>Missing: 36/1569 [2.2%] |
| BMI on Enlistment | Median: 22.3<br>Min: 16.7<br>Max: 37.6 |
| Measles Hospitalisation | Yes: 65/1569 [4.1%]<br>No: 1504/1569 [95.9%] |
| Measles (n = 65) | Earliest: 25 May 1915 |

Finally, to assess the reproducibility of the database, data extraction was repeated by an independent investigator for 100 randomly chosen individuals. Compared to the original database, a 0.12% error rate was noted in the re-recorded data. The low error rate recorded here suggests strong reproducibility in data extraction.

### Military personnel characteristics

Having validated key aspects of the database we next sought to describe the population cohort (Table 1). The median age of the population was 23 (min: 15, max: 52) and 99.8% of individuals were male (Table 1). 81.7% of individuals held a blue-collar job prior to enlistment and 48.3% were born in urban areas (Table 1). Descriptions of complexion varied significantly and often appeared somewhat arbitrary (Supplementary Table 2) although almost 60% of individuals were classified as having a ‘light’ complexion (Table 1). Approximately 4% of individuals had a recorded hospitalisation with measles during the study period, with the first hospitalisation recorded on the 25^th^ May 1915 and the last hospitalisation recorded on the 25^th^ of March 1919 (Table 1). Significant force-dependent differences were observed in the cohort. Namely, those from CEF were significantly more likely to survive the duration of the study period, experience reduced pandemic ILI severity and have the highest rate of hospitalisation for measles (Figure 1E). Of those who were recorded as deceased during the study period, the majority (75%) died on the Western Front or in England.

### Measles increases the risk of secondary infections, excluding pandemic ILI

Prior to the availability of the measles vaccine in the 1960s measles was responsible for significant mortality in the armed forces^6,7,9^. Consistent with these data, of the 65 members of the AIF, NZEF and CEF who were hospitalised for measles during the study period, approximately 6% died directly of the infection (Figure 2). For individuals who survived the initial infection, 32% of individuals died of an infectious disease (Figure 2).

**Figure 2:**
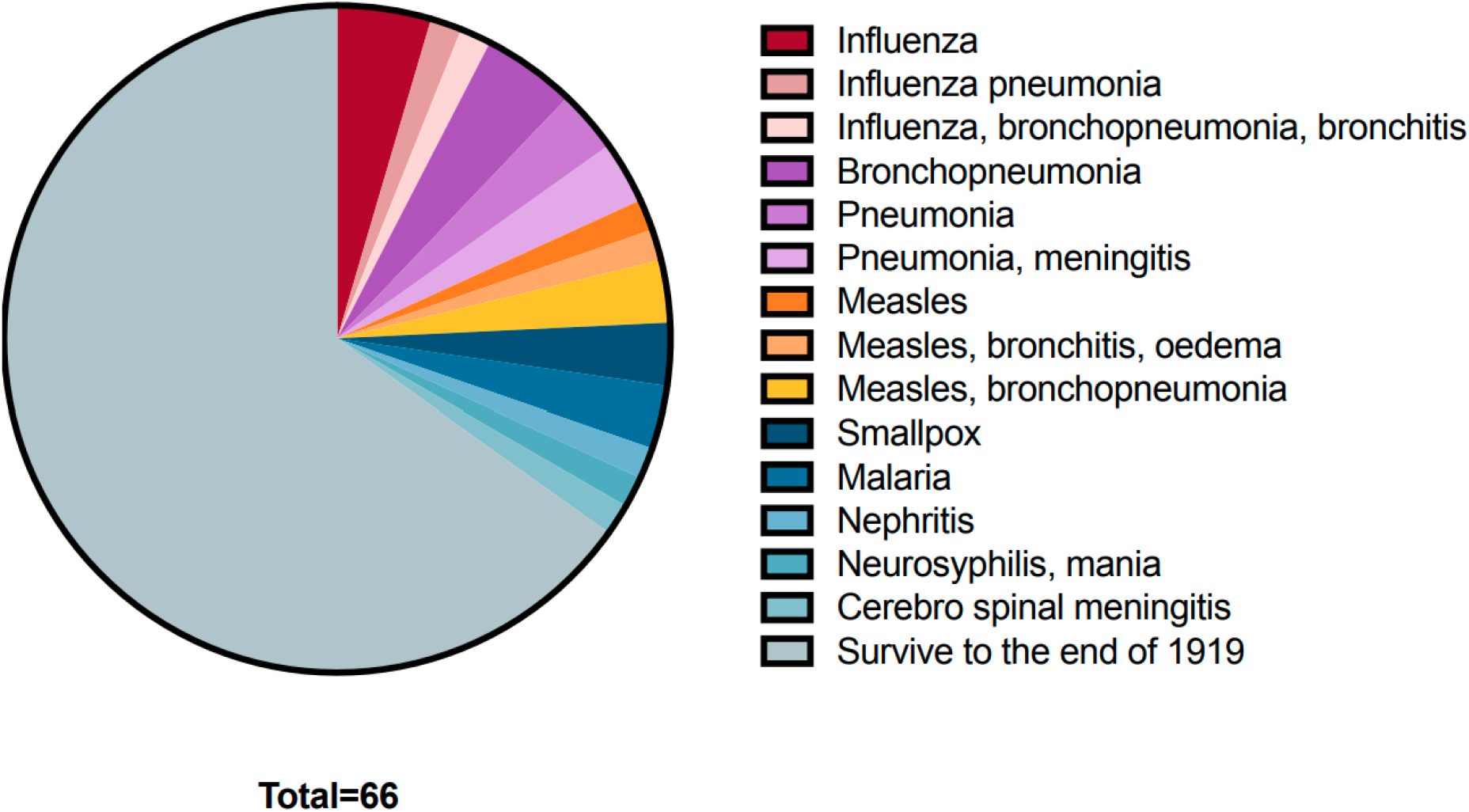
Primary cause of death in military personnel with a history of measles hospitalization during the study period.

To further assess the impact of a recent measles hospitalisation on the severity of non-pandemic ILI infection a multiple logistic regression model was run using predictor variables measles (yes/no), force and age at time of enlistment. In this analysis we define the binary outcome 1 = “death by infection (excluding pandemic ILI)” and 0 = “no death, death by non-infection, or death by pandemic ILI”. Table 2 shows that a recent history of measles statistically significantly increases the probability of an individual’s classification into outcome 1 (p = 0.03) [i.e. death by infection (excluding pandemic ILI)] after age and force are controlled in the model. Taken together, these data show the significant direct and indirect detrimental effects that measles had on individuals of the AIF, NZEF and CEF during WW1.

**Table 2:**
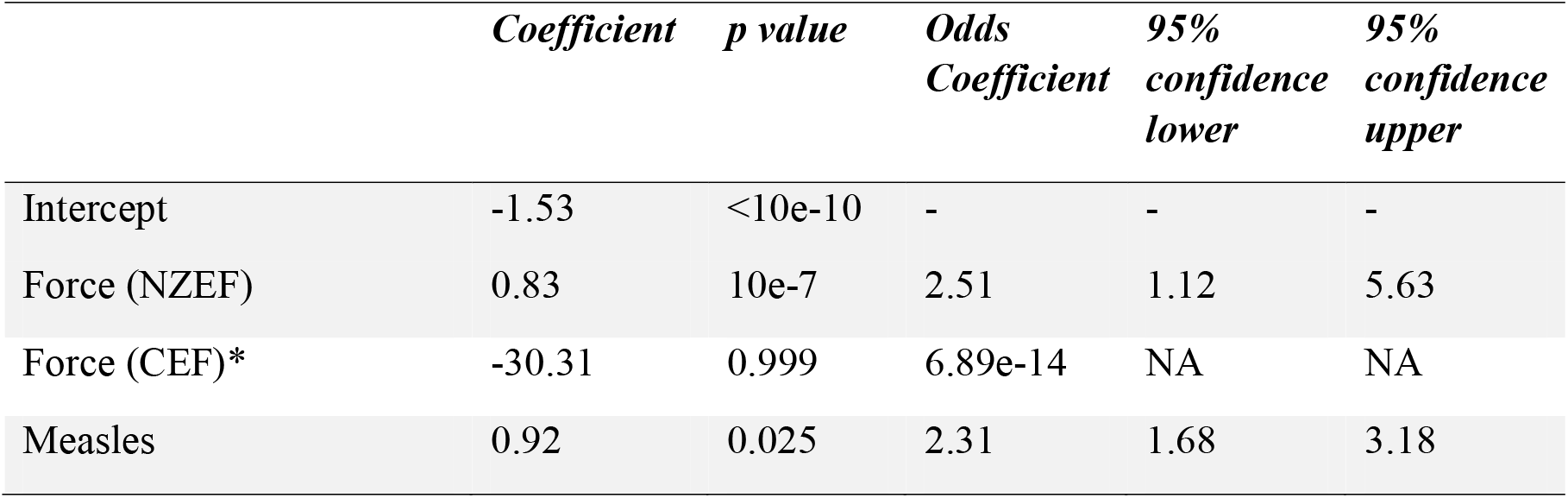
Regression coefficients and p-values from multiple logistic regression of death by infectious disease excluding pandemic ILI. *No death by infectious disease excluding pandemic ILI was observed in the Canadian force.

|  | *Coefficient* | *p value* | *Odds* | *95%* | *95%* |
| --- | --- | --- | --- | --- | --- |
|  |  |  | *Coefficient* | *confidence* | *confidence* |
|  |  |  |  | *lower* | *upper* |
| Intercept | -1.53 | <10e-10 | - | - | - |
| Force (NZEF) | 0.83 | 10e-7 | 2.51 | 1.12 | 5.63 |
| Force (CEF)* | -30.31 | 0.999 | 6.89e-14 | NA | NA |
| Measles | 0.92 | 0.025 | 2.31 | 1.68 | 3.18 |

### Prior measles infection and the severity of pandemic ILI

To assess if a prior measles infection had a similar effect on pandemic ILI as observed for other infectious disease, multiple linear regression was used to assess the number of days of hospitalisation for pandemic ILI controlling for force, age, occupation, complexion, BMI, prior hospitalisation for seasonal ILI and prior hospitalisation for measles (Table 3). Age showed a significant relationship with days hospitalised for pandemic ILI, with older individuals being hospitalised for fewer days (p = 0.015). Prior hospitalisation with seasonal ILI was association with increased duration in hospitalisation for pandemic ILI (p = 0.015) whilst a prior measles infection was associated with reduced time in hospital from pandemic ILI (p = 0.037).

**Table 3:** Regression coefficients and p-values from multiple linear regression of days in hospitalisation for pandemic ILI in individuals of the AIF, AZEF and CEF during WW1^1^.

|  | coefficient | p value |
| --- | --- | --- |
| Intercept | 10.94 | 0.029 |
| Force (NZEf) | 0.63 | 0.258 |
| Force (CEF) | 1.43 | 0.607 |
| Age | -0.18 | 0.015 |
| Occupation (White collar) | 0.43 | 0.688 |
| Occupation (Mixed/Undefined) | -7.80 | 0.589 |
| Complexion Code | 0.91 | 0.272 |
| BMI | 0.09 | 0.629 |
| Prior hospitalisation for seasonal ILI | 3.91 | 0.015 |
| Prior hospitalisation for mustard gas | -1.02 | 0.534 |
| Prior hospitalisation for measles | -4.41 | 0.037 |

## DISCUSSION

Here, we showed the devasting effects that measles virus infections had on military populations in WW1. Mortality for measles could be as high as 12% in certain military populations^6^ and in the present study we recorded a 6% measles mortality rate amongst those who were hospitalised with the virus. A wealth of recent data indicates that measles also has detrimental effects on those individuals who are fortunate enough to survive the initial acute infection. Specifically, measles virus can infect memory B and T cells, resulting in immune amnesia for up to three years after the resolution of the acute infection^1,2^. Accordingly, in modern times incomplete measles vaccination coverage and subsequent measles virus infection is associated with an increased susceptibility to a variety of other infectious diseases^4^. Here, we show the same association in WW1, which occurred approximately 45 years prior to the development of the measles vaccine. Although our evidence for this association between a prior measles virus infection and the development of a secondary infection is only correlative, it is consistent with our modern understanding of measles virus pathogenesis.

Based on the aforementioned studies and data it is tempting to speculate that a prior infection with measles virus resulted in an ablation of pre-existing influenza virus immunity and the increased severity of 1918 pandemic ILI^6,25^. Strikingly, the present study found just the opposite: when controlling for other key co-variates a prior measles hospitalisation was associated with a reduced severity of pandemic influenza. This is consistent with prior data, which showed that areas of Newfoundland, Canada that experienced widespread measles outbreaks in 1916 and 1917 were largely unaffected by pandemic influenza in 1918^26^. This is despite the widespread mortality from the influenza pandemic in adjacent communities^26^. It is interesting to speculate the mechanisms by which this may occur. It has previously been suggested that pro-inflammatory T cells played a significant role in the pulmonary pathology associated with 1918 influenza^22^. Accordingly, in this unique context the depletion of T cells by measles virus may be beneficial in terms of influenza severity. Alternatively, it is possible these data reflect the phenomenon of original antigenic sin. Original antigenic sin refers to the fact that after a person’s first influenza infection, their immune system tends to recall and reuse memory cells against that initial strain during later infections. As a result, antibody responses are directed toward the original virus rather than the new strain’s unique epitopes, which can lead to reduced protection against novel influenza variants. If previous influenza virus strains were markedly different to that of the pandemic 1918 virus, original antigenic sin could mean that the depletion of prior immunity could have been beneficial in the generation of 1918 specific immune responses. Establishing a true causal relationship between a prior measles virus infection and reducing 1918 influenza severity is an important goal for future studies. Importantly, ferret models for the 1918 influenza virus^36^ and co-infections with morbilliviruses and influenza virus^37^ have already been established. Such models could represent an invaluable tool in understanding the true relationship between a prior measles virus infection and 1918 influenza severity.

In addition to measles, the present study also provided important insights into other factors associated with hospitalisation duration of 1918 ILI amongst military populations. Consistent with what has been described previously^22^, younger age was associated increased time in hospital for pandemic ILI – reflecting the unique demography of the 1918 pandemic. Interesting, prior hospitalisation with seasonal ILI was associated with increased hospitalisation duration for 1918 ILI. This may once again reflect original antigenic sin. Had seasonal influenza (which assumedly was very antigenically distinct from the pandemic virus) been an individual’s first influenza virus infection, it is possible that during the 1918 pandemic the majority of the antibody response was directed against seasonal virus epitopes and hence not necessarily protective in the pandemic context.

In our multiple linear regression of days in hospitalisation for pandemic ILI military force did not have a significant effect. However, we showed clearly that members of the CEF were significantly more likely to survive the study period than individuals in the AIF or NZEF. These data may reflect the different search strategy used for individuals in the CEF. Alternatively, it is possible that these data reflect a real historical phenomenon where Canadians faired better in WW1 than other members of the allied forces. There is scarce historical data available that describes the day-to-day conditions of the CEF compared to either the AIF or NZEF. It is possible that these data reflect better healthcare amongst the CEF or different locations of deployment, but such stark difference in mortality warrants further investigation.

The searchable database developed herein can be used to answer a wide variety of other research questions in the future. Military populations represent a unique resource of information on diseases in WWI due to the meticulous and extensive records that were kept. Indeed, such records have already improved our understanding of 1918 influenza^29-31^. However, unlike previous studies the database developed herein combined the records of three Imperial/Expeditionary forces and documented all infectious disease hospitalisation from 1915 onwards. In the future it is hoped that advances in artificial intelligence (AI) will facilitate the more high-throughput screening of scanned military records and enable the creation of yet larger, searchable, datasets. Nevertheless, the validation of our data collection methods as well as the analysis of the collected data suggested that even in the absence of AI, minimal errors occurred in database creation and this database can be used to interrogate other aspects of health in WWI.

The present study was subject to several limitations. Firstly, whilst > 300,000 records in total were screened only 1,569 individuals met the pre-defined study criteria, of which only 65 recorded a measles hospitalisation during the study. This limited n number restricted the type of analysis that could be performed. Nevertheless, the use of this sample size was justified by our power analysis and the fact that other well known contributors to the severity of 1918 influenza (e.g. age) were also identified in our analysis. Furthermore, it must be recognised that the relationships observed herein were not necessarily causative and these data should be interpreted as an impetus for further experimental research into the interactions between measles and pandemic viruses. Finally, we acknowledge that this analysis is based on the assumption that all ILI in 1918 were indeed caused by pandemic influenza, although its possible that other infections (e.g. RSV or bacterial infections) contributed to ILI presentations. Despite these limitations, the present study highlights yet another unique feature of the 1918 influenza pandemic: unlike other infectious diseases, prior hospitalisation with measles was associated with a reduction in 1918 pandemic severity. However, the high mortality rate caused by measles itself and the increased risk for severe infections, other than the 1918 pandemic virus, remain important reasons to encourage measles vaccination.

## Data Availability

All data produced in the present study are available upon reasonable request to the authors

## ACKNOWLEDGEMENTS

We would like to thank Kandace Bogaert for provision of data for the CEF and Dennis Shanks for informative discussions. We are truly thankfully for the data made available from all three forces and we would like to acknowledge the sacrifices made by all the individuals included in this study. We would also like to thank CAS and the Norwegian Academy of Science to work make this work possible, Kirsty R. Short is supported by an NHMRC investigator grant 2007919. CES is supported by the ARC-DECRA Fellowship (#DE220100185) and the University of Melbourne Establishment Grant.

## Footnotes

1 Library and Archives of Canada, RG 9 II-L-1, Volumes 1-4, 6: Military Hospital, London, Ontario (books 1-3, 5-11), Wolseley Barracks Hospital, London, Ontario (books 12-20), Overseas men in Civil Hospital, London, Ontario (book 22), Convalescent Military Hospital, London, Ontario (books 23-31), Officers and Men in Civil Hospital (book 32), Victoria Hospital, London, Ontario (book 33), Royal Naval Canadian Volunteer Reserve Hospital Officers and Nursing Sisters Individuals Dependants, London, Ontario (book 34), Canada Cases Officers Nursing Sisters (N/S) and Pensioners, London, Ontario (book 35), Lord Dufferin Hospital, Orangeville, Ontario (book 36), Oshawa General Hospital, Oshawa, Ontario (book 37), Wingham General Hospital, Wingham, Ontario (book 38), Canada Camp Cases Officers N/S and Pensioners Imperial Army, London, Ontario (book 39), County of Bruce General Hospital, Walkerton, Ontario (book 40), General Hospital, Guelph, Ontario (book 41), Guelph Military Hospital, Guelph, Ontario (book 42), Galt General Hospital, Galt, Ontario (book 43), Canadian Army Medical Corps Hospital, London, Ontario (books 44-48), Hotel Dieu Hospital, Windsor, Ontario (book 48A), Military Base Hospital, Toronto, Ontario (book 81), Base Hospital, Toronto, Ontario (book 81B), Whitby Military Hospital, Whitby, Ontario (books 82-95), St Andrews Hospital, Toronto, Ontario (books 96-107, 110-111), Women’s Aid Department, St Andrews Hospital, Toronto, Ontario (book 108), Military Hospital, Hamilton, Ontario (books 112-119), St Josephs Military Hospital, Hamilton, Ontario (book 120), Niagara Camp Hospital, Niagara (books 121-122, 125A), Niagara General Hospital, Niagara Falls (book 125B), Lady Minto Hospital, Chapleau, Ontario (book 126), Lady Minto Hospital, New Liskeard, Ontario (book 127), Lady Minto Hospital, Cochrane, Ontario (book 128), Spidina [sic] Military Hospital, Spidina [sic], Ontario (books 129-133), General Hospital, Sault Ste Marie, Ontario (book 173), General Hospital, Parry Sound, Ontario (book 174), Providence General Hospital, Haileybury, Ontario (book 175), Kaipuskasing [sic] Station Hospital, Ontario (book 181), Welland County Hospital, Welland, Ontario (book 182), Special Hospital Royal Air Force (books 183-184), Exhibition Camp Hospital, Toronto, Ontario (books 185-187), Exhibition Camp Hospital, Niagara (book 187D), Exhibition Camp, Toronto, Ontario (book 187E), General Hospital, Orillia, Ontario (book 188), Queen Victoria Memorial Hospital, North Bay, Ontario (book 189), Euclid Hall Hospital, 515 Jarvis Street, Toronto, Ontario (book 190), Queens Military Hospital, Kingston, Ontario (books 191, 194-196, 198-200), and Canadian General Stationary Hospital, Hamilton, Ontario (book 193)

